# Incidence of Right Ventricular Dysfunction in an Echocardiographic Referral Cohort

**DOI:** 10.1101/2024.10.08.24315120

**Authors:** Jonah D. Garry, Shi Huang, Jeffrey Annis, Suman Kundu, Anna Hemnes, Matthew Freiberg, Evan L. Brittain

## Abstract

**Introduction:** Incidence rates (IRs) of RV dysfunction (RVD) are unknown. We examined the rates, risk factors, and heart failure (HF) hospitalization hazard associated with incident RVD in patients referred for Transthoracic Echocardiogram (TTE).

**Methods:** In this retrospective cohort study, we extracted tricuspid regurgitant velocity (TRV) and tricuspid annular systolic plane excursion (TAPSE) from TTEs at Vanderbilt (2010-2023). We followed patients from their earliest TTE with normal RV function (TAPSE≥17mm) and a reported TRV. The primary outcome was new RVD (TAPSE<17mm), and the secondary outcome was HF hospitalization after second TTE. Poisson regression and multivariable cox models estimated IRs and hazard ratios, adjusted for demographics, comorbidities, and TTE measures.

**Results:** Among 45,753 patients (63 years [IQR 50-72], 45% Male, 13% Black) meeting inclusion criteria, 13,735 (30.1%) underwent a follow up TTE and 4,198 (9.2%) developed RVD. The IR of RVD in the full cohort was 3.2/100 person/years (95%CI 3.1-3.3) and 8.2 (95%CI 8.0-8.5) in the repeat TTE cohort. IRs increased with rising RVSP. Risk factors for incident RVD were most prominently HF (HR 1.88; 95%CI 1.75–2.03), left-sided valvular disease (HR 1.68; 95%CI 1.53–1.85), and other cardiovascular comorbidities. Baseline RVSP >35 mmHg associated with TAPSE decline over time. Incident RVD increased hazard of HF hospitalization (HR 2.02; 95%CI 1.85-2.21). Hazard of HF hospitalization increased when TAPSE declined by ≥5mm.

**Conclusions:** RVD incidence is substantial among patients referred for TTE. Clinical monitoring is warranted if RVSP >35mmHg. Cardiovascular comorbidities drive RVD in this population. Incident RVD associates with increased hazard of HF hospitalization.

## Introduction

Right ventricular dysfunction (RVD) is associated with increased morbidity and mortality in the general population^1^ and across the spectrum of cardiovascular disease,^2–7^ however there are substantial gaps in current understandings of RV epidemiology.^8^ To date, studies of RVD have focused on prevalence and prognostic implications in highly selected populations, with small studies examining the impact of RVD within a specific underlying etiology such as heart failure (HF) with further subdivision by left ventricular ejection fraction (LVEF)^9–12^ and pulmonary hypertension (PH)^5^ with stratification by WSPH group or etiology.^13–15^ The few population-level epidemiologic investigations of RVD to date derive from the NHLBI cohorts MESA,^16^ ARIC,^17^ and CARDIA^18^ and provide cross-sectional prevalence using multiple imaging metrices of RV function. No studies to date have described the incidence rate (IR) of RVD longitudinally or evaluated the clinical characteristics associated with development of RVD in a large-scale population of patients seeking care.

Here, we report the first incidence rates of RVD among patients referred for echocardiography and identify the associated clinical risk factors to better understand population-level drivers of RVD. We hypothesized that the predominant factors associated with RVD would be cardiovascular comorbidities in addition to pulmonary pressure and that incident RVD would be associated with an increased hazard of heart failure hospitalization.

## Study Design and Methods

We conducted a retrospective cohort study using deidentified electronic medical record data from a tertiary care center. The local Institutional Review Board approved the study. We extracted TTE reports from the electronic health record as previously described.^19,20^ We assessed all adult patients (aged ≥ 18 years) referred for TTE between May 7, 2010 (when TTE reports began recording Tricuspid Annular Plane Systolic Excursion [TAPSE)) and August 2023.

### Exposures, Covariates, and Outcomes

We selected the first TTE available with normal RV function (TAPSE ≥17mm) and a concurrently available TRV for each patient as the baseline. Overall TRV availability (46.3%) was comparable to the similarly sized contemporaneous National Echo Database Australia cohort and higher than the CARDIA echocardiography core laboratory.^21–23^ Patients who had a TTE with TAPSE <17mm before a subsequent TTE with normal RV function were excluded. The predictor of interest was PH, defined by a TRV >2.8m/s based on the 2022 ERS/ERC guidleines.^24^ The primary outcome of interest was the development of RVD on a follow up TTE defined as TAPSE <17mm.^25^ Routine TAPSE measures were instituted in our echocardiography lab in 2011 after publication of the ASE guidelines for assessment of the right heart^26^ and a contemporaneous primer for cardiac sonographers^27^ following a strict quality control period. TAPSE measures were generated by RDCS certified sonographers and overread by attending echocardiographers. We identified the temporally closest follow-up TTE demonstrating RVD. For patients who did not develop RVD, we censored patients at the date of last medical contact or death. We excluded patients with fewer than 3 months follow-up to avoid biasing incident RVD estimates with acute changes to RVD.

We abstracted demographic, laboratory, echocardiographic, and comorbidity information indexed to the date of baseline TTE in a similar fashion to prior studies.^28–30^ Further details regarding data abstraction are included in the ***Supplemental Methods*.**

### Incidence Rate Analyses

We estimated IRs for the overall cohort, stratified by PH status, and stratified by RVSP categories. Our primary IR estimates using the Full Cohort assumed conservatively that all patients without a second TAPSE measurement did not develop RVD (“minimum estimate”). We then performed secondary analysis limiting the analysis cohort to patients with at least one follow up TAPSE measurement (“Repeat TTE Cohort”), using the temporally closest TTE and derived a “maximum estimate.” This dual approach provides maximum information from our cohort by offering upper and lower bounds for RVD IR estimates. To assess whether long-term follow up without repeat TTE could bias our results to lower rates we also performed a sensitivity analysis of the full cohort censoring all patients at 5-years of follow up.

### Characteristics Associated with RVD and TAPSE

We assessed the association between baseline characteristics and development of RVD in the Full Cohort and Repeat TTE Cohort. In secondary analyses we re-assessed these associations stratified by PH status. We performed sensitivity analyses with time-varying comorbidities to assess whether new diagnoses explained some degree of incident RVD hazard. Details of exposure time, follow up echo selection, and variable selection for these models are noted in the ***Supplemental Methods***.

### HF Hospitalization

We conducted a time-to-event analysis to assess the impact of incident RVD and change in TAPSE between TTEs on the hazard of HF hospitalization or rehospitalization. We performed a sensitivity analysis of only those patients with their second TTE conducted outpatient with inpatient TTE status determined the presence of any inpatient ICD code within one day of the TTE. Details of HF hospitalization definition, exposure time, and inpatient/outpatient TTE status are detailed in the ***Supplemental Methods*.**

### Right Ventricle-Pulmonary Artery (RV-PA) Uncoupling

In secondary analysis, we assessed the IRs, risk factors, and HF hospitalization hazard associated with incident RV-PA Uncoupling. We defined RV-PA Uncoupling by TAPSE/RVSP <0.36 mm/mmHg based on prior reports.^5,31^ For this analysis we excluded patients in the full cohort with prevalent RV-PA uncoupling and included follow up TTEs with both TAPSE and RVSP available.

### Statistical Analysis

Patient characteristics were described using frequency with percentages for categorical variables and median with interquartile range for continuous variables. We assessed differences in clinical characteristics by PH status using Pearson chi-squared testing for categorical variables and Wilcoxon-Rank Sum testing for continuous variables. IRs for RVD were calculated as cases per 100 person-years, with estimated 95% confidence intervals generated under Poisson distribution assumption and by exact method. We used Kaplan Meier curves to display the time to development of RVD with log-rank testing to compare between patients with and without PH. We examined the relationship between baseline characteristics and hazard of developing RVD using multivariable cox regression, adjusting for demographics, comorbidities, and baseline echo measures. We assessed the relationship between baseline characteristics and change in TAPSE using Ordinary Least Squares regression, adjusting for baseline TAPSE and length of follow up time. In all multivariable models, the effects of continuous predictors were assumed nonlinear, using restricted cubic splines with three knots. We report predictors’ effect graphically and by comparing the 75^th^ percentile value to the 25^th^ percentile value. We assessed the relationship between incident RVD/change in TAPSE and HF hospitalization using multivariable Cox regression and adjusting for demographics, time-varying comorbidities, and baseline TTE measures.

All models were conducted using complete case analysis. We excluded baseline variables with >10% missingness in the primary regression models. Statistical Analyses were preformed using R software (version 4.3.1; www.r-project.org).

## Results

We identified 45,753 patients without RVD who had a baseline TTE with TRV and TAPSE measurements available and met our inclusion criteria (Full Cohort; **Figure 1**). Of these, 9,700 patients had PH (PH+ cohort) and 36,053 did not (PH-cohort). Baseline characteristics stratified by PH status are displayed in **Table 1**. There were 13,735 patients referred for a follow up TTE over the study period (Repeat TTE Cohort). **eTable 1** displays characteristics compared between the Full Cohort and Repeat TTE Cohort.

**Figure 1:**
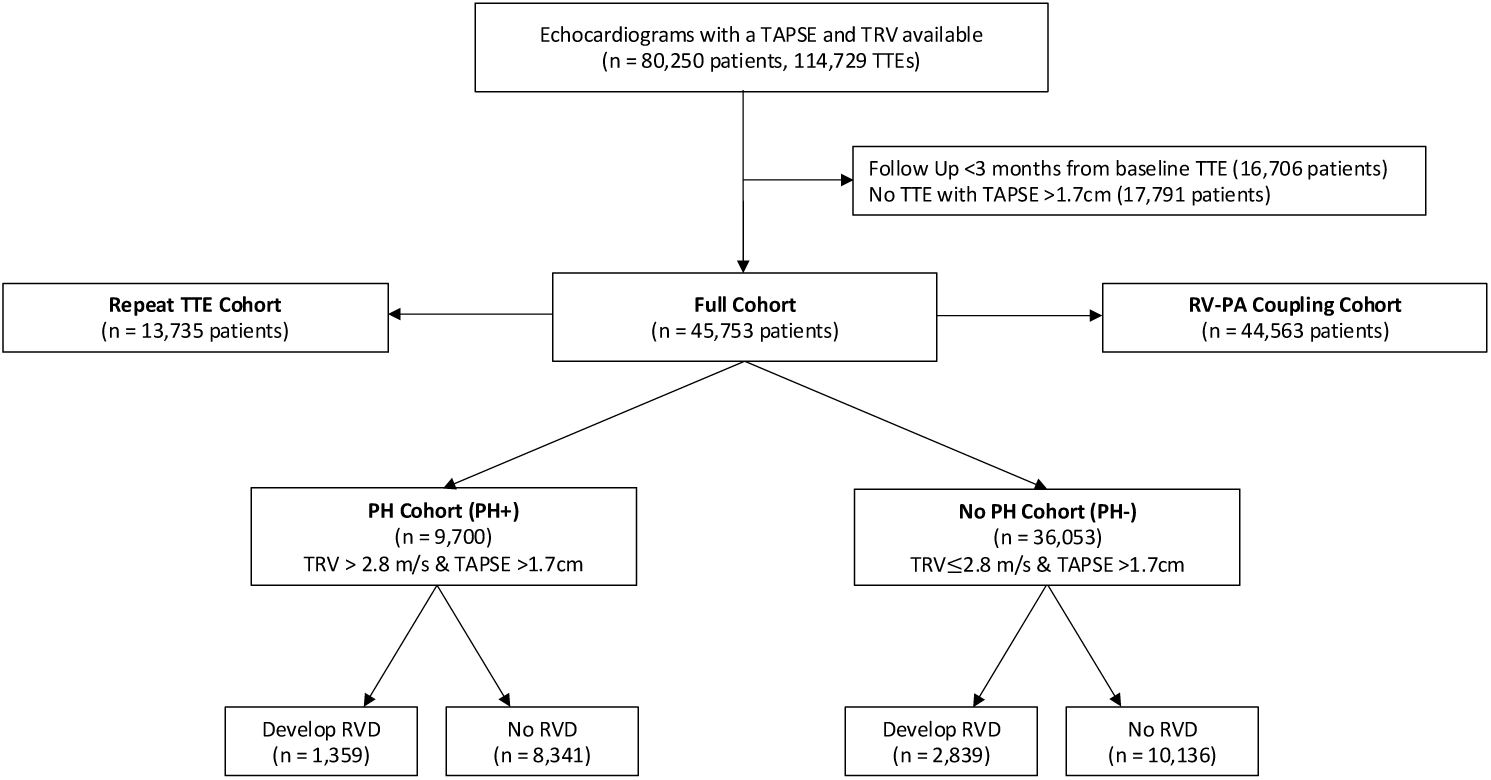
Cohort Identification Diagram. Figure 1 displays the process of identification of the study cohort and reasons for echocardiogram/patient exclusion

**Table 1:** Clinical Characteristics at Baseline Transthoracic Echocardiogram by PH status (Full Cohort n = 45753)

| Characteristic | PH absent (PH-)<br>n = 36053 | PH present (PH+)<br>n = 9700 | p-value |
| --- | --- | --- | --- |
| Age in years | 61 (47–71) | 68 (58–77) | <0.001 |
| Male Sex | 16410 (46%) | 4309 (44%) | 0.055 |
| BMI in kg/m<sup>2</sup> | 28.7 (24.8–33.6) | 29.2 (25.1–34.8) | 0.001 |
| Race/Ethnicity |  |  | 0.001 |
| White, Non-Hispanic | 29124 (81%) | 7551 (78%) |  |
| Black | 4175 (12%) | 1589 (16%) |  |
| Hispanic | 529 (1%) | 121 (1%) |  |
| Other | 1274 (4%) | 278 (3%) |  |
| Baseline TAPSE in cm | 2.20 (1.99–2.50) | 2.20 (1.95–2.56) | 0.680 |
| Baseline RVSP in mmHg | 26 (22–31) | 44 (39–52) | <0.001 |
| LV Ejection Fraction | 62 (56–67) | 61 (55–67) | <0.001 |
| Left-Sided Valvular Disease | 1183 (3%) | 914 (9%) | <0.001 |
| Tricuspid Regurgitation | 243 (1%) | 620 (6%) | <0.001 |
| Left Atrial Diameter in cm | 3.7 (3.2–4.1) | 4.1 (3.6–4.6) | <0.001 |
| Average E/e' | 8.3 (6.5–10.6) | 10.9 (8.1–14.8) | <0.001 |
| Diastolic Function |  |  | <0.001 |
| Normal | 7952 (22%) | 1089 (11%) |  |
| Grade 1 | 5328 (15%) | 1233 (13%) |  |
| Grade 2 | 680 (2%) | 643 (7%) |  |
| Grade 3 | 119 (1%) | 235 (2%) |  |
| eGFR in mL/min/1.73m<sup>2</sup> | 80 (63–96) | 68 (45–87) | <0.001 |
| SBP in mmHg | 126 (115–140) | 130 (117–146) | <0.001 |
| Systemic Sclerosis | 217 (1%) | 109 (1%) | <0.001 |
| Heart Failure | 7783 (22%) | 4406 (45%) | <0.001 |
| Atrial fibrillation | 7595 (21%) | 3495 (36%) | <0.001 |
| Coronary Artery Disease | 9855 (27%) | 3739 (39%) | <0.001 |
| Interstitial Lung Disease | 4249 (12%) | 2086 (22%) | <0.001 |
| COPD | 3762 (10%) | 2027 (21%) | <0.001 |
| Obstructive Sleep Apnea | 6636 (18%) | 2253 (23%) | <0.001 |
| HIV | 392 (1%) | 108 (1%) | 0.830 |
| Diabetes Mellitus | 8479 (24%) | 3473 (36%) | <0.001 |
| Hyperlipidemia | 19756 (55%) | 6211 (64%) | <0.001 |
| Follow Up in days | 794 (368–1562) | 625 (297–1288) | <0.001 |
| Variable Missingness in Overall Cohort: Race/Ethnicity: 1112 (2.4%); BMI: 828 (1.8%); eGFR: 11302 (24.7%); SBP 492 (1.1%); LVEF 490 (n=1.1%); LA diameter 8022 (17.5%); Average LV E/e' ratio (67.3%); Diastolic Function 28,474 (62.2%) |  |  |  |

The PH+ cohort expectedly had a higher RVSP (44 mmHg vs 26 mmHg) and were older (68 vs 61 years), with similar sex distribution, body mass index (BMI), and racial/ethnic composition. Baseline TAPSE was similar between the two groups (2.20 vs 2.20, p = 0.68). There was a higher prevalence of cardiometabolic and pulmonary comorbidities in the PH+ cohort including HF (45% vs 22%) and AF (36% vs 21%) as well as a higher prevalence of cardiac structural pathology. Median follow up was shorter in the PH+ cohort (625 days [IQR 297-1288] vs 794 days [IQR 368-1562]). Ultimately 1690 patients (4.7%) without PH at baseline newly developed PH on their follow up TTE.

### Incidence Rates of Right Ventricular Dysfunction

The primary outcome (new RVD) occurred in 4,198 patients (10.1%). The median time to development of new RVD was 765 days (IQR 364-1466), while patients who did not develop RVD (followed until death or last medical contact) were followed for a median of 758 days (IQR 353-1045). For patients with a second TAPSE measurement, the mean change in TAPSE was - 2.44mm +/− 5.78mm. Unsurprisingly, there was a larger TAPSE decline among the subset of these patients who developed incident RVD (−7.18mm +/− 4.37) compared to patients who did not (−0.52mm +/− 5.14). The primary outcome occurred in 14.0% of patients with PH (n = 1,359 patients) and 7.9% of patients without PH (n=2,839). Of note, the proportion of patients with a repeat TTE was different between the PH+ and PH-cohorts (37.1% vs 28.1%).

The IR estimate of RVD in the Full Cohort (minimum estimate) was 3.2 per 100 person-years (95%CI 3.1–3.3), reaching up to 8.2 per 100 person-years (95%CI 8.0–8.5) in the Repeat TTE cohort (maximum estimate). Rates were nearly identical with censoring at 5-years of follow up. IRs of RVD in the PH+ cohort were 1.6-2.1x higher than the PH-cohort. IRs of RVD increased consistently as RVSP increased (**Table 2**). Kaplan-Meier curves for the development of RVD stratified by PH status are displayed in **Figure 2**.

**Figure 2:**
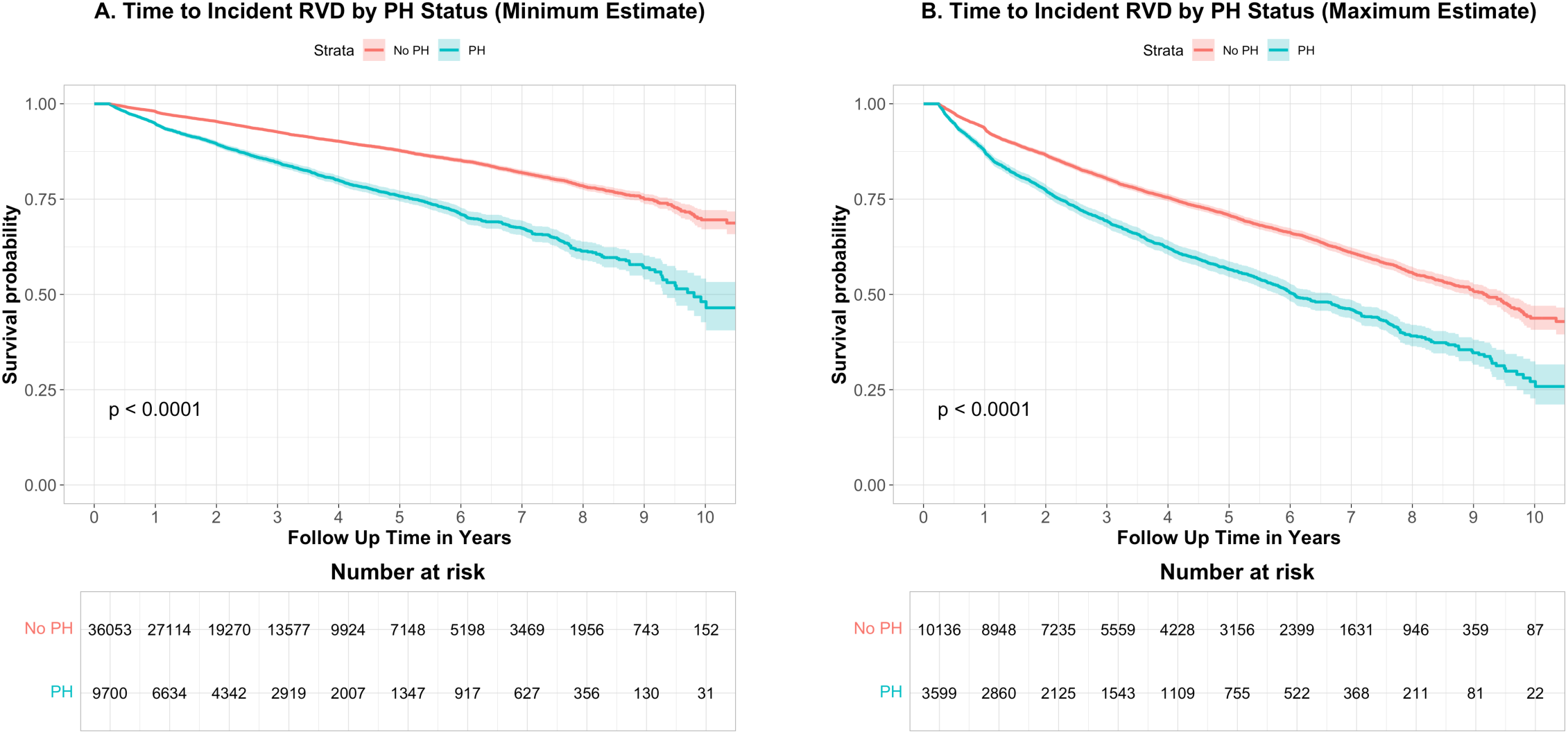
Kaplan Meier Curves for Incidence of Right Ventricular Dysfunction. Figure 2 displays Kaplan Meier Curves for the Incidence of Right Ventricular Dysfunction in the Full Cohort (A), representing a minimum estimate, and in the Repeat TTE Cohort (B), representing a maximum estimate.

**Table 2:** Incidence Rate Estimates of Right Ventricular Dysfunction and RV-PA Uncoupling.

|  | **TAPSE** |  | **TAPSE/RVSP Ratio** |  |
| --- | --- | --- | --- | --- |
|  | **Full cohort**<br>(Minimum estimate) | **Repeat TTE Cohort**<br>(Maximum estimate) | **Full Cohort**<br>(Minimum estimate) | **Repeat TTE Cohort**<br>(Maximum estimate) |
|  | **Rate/100 PY (95% CI)** |  |  |  |
| **Entire follow up** | 3.2 (3.1 – 3.3) | 8.2 (8.0 – 8.5) | 1.2 (1.1 – 1.3) | 3.4 (3.2 – 3.6) |
| **5-year Limited** | 3.1 (3.0 – 3.2) | 8.2 (7.9 – 8.4) | 1.1 (1.0 – 1.2) | 3.3 (3.1 – 3.5) |
| **Stratified by Baseline PH Status** |  |  |  |  |
| **PH +** | 5.6 (5.3 – 5.9) | 11.9 (11.3 – 12.6) | 3.6 (3.3 – 3.8) | 9.3 (7.7 – 8.9) |
| **PH -** | 2.7 (2.6 – 2.8) | 7.2 (6.9 – 7.5) | 0.7 (0.7 – 0.8) | 2.2 (2.1 – 2.4) |
| **Stratified by RVSP Category** |  |  |  |  |
| **< 30 mmHg** | 2.5 (2.3 – 2.6) | 6.9 (6.6 – 7.2) | 0.5 (0.5 – 0.6) | 1.5 (1.3 – 1.6) |
| **[30-40) mmHg** | 3.3 (3.2 – 3.5) | 8.1 (7.6 – 8.6) | 1.5 (1.4 – 1.6) | 3.5 (3.2 – 3.9) |
| **[40-50) mmHg** | 5.4 (5.0 – 5.9) | 11.7 (10.8 – 12.7) | 4.0 (3.6 – 4.5) | 8.5 (7.6 – 9.4) |
| **[50-60) mmHg** | 6.5 (5.7 – 7.4) | 13.1 (11.5 – 14.9) | 5.1 (4.2 – 6.1) | 10.6 (8.7 – 12.7) |
| **>60 mmHg** | 9.9 (8.8 – 11.1) | 17.3 (15.3 – 19.4) | 6.2 (4.3 – 8.6) | 14.0 (9.8 – 19.5) |
| NOTE: Percentage of patients with a follow up TTE varies between cohorts which may impact upper bound estimate. |  |  |  |  |

### Baseline Characteristics Associated with Incident RVD

The baseline characteristics most strongly associated with RVD in multivariable regression modeling were HF (HR 1.88; 95%CI 1.75-2.03), systemic sclerosis (HR 1.84; 95%CI 1.41–2.39), atrial fibrillation (AF) (HR 1.54; 95%CI 1.44–1.65), and left sided valvular disease (HR 1.68; 95%CI 1.53–1.85). The full regression model is displayed in **eTable 2**. Notably, coronary artery disease (CAD), diabetes, interstitial lung disease (ILD), and Male Sex were associated with a significantly increased hazard of RVD while moderate or worse tricuspid regurgitation was associated with a decreased hazard of RVD (HR 0.84 [95%CI 0.72–0.98]). Associations between continuous variables and probability of RVD at 3 years (median follow-up) are displayed in **Figure 3**. TAPSE, BMI, and LVEF were non-linearly associated with RVD (all p<0.001). The probability of RVD at 3 years decreased steeply as baseline TAPSE increased from 1.7 until 2.5, above which the probability of RVD continued to decrease more gradually. The probability of RVD is consistently highest for BMIs <30, however as BMI increases above 30 the probability of RVD steadily decreases. For LVEF the probability of RVD appears to linearly decrease from the lowest values up to 60, above which there is a small increase in probability of RVD. The probability of RVD decreased linearly as age and eGFR increased and increased linearly as RVSP and LA diameter increased (all p<0.05). Higher stages of diastolic dysfunction were associated with increased hazard of RVD, while LV average E/e’ was not significantly associated with hazard of RVD. Addition of eGFR and diastolic function to the model did not significantly change the association of any other variables, including HF or LVEF.

**Figure 3:**
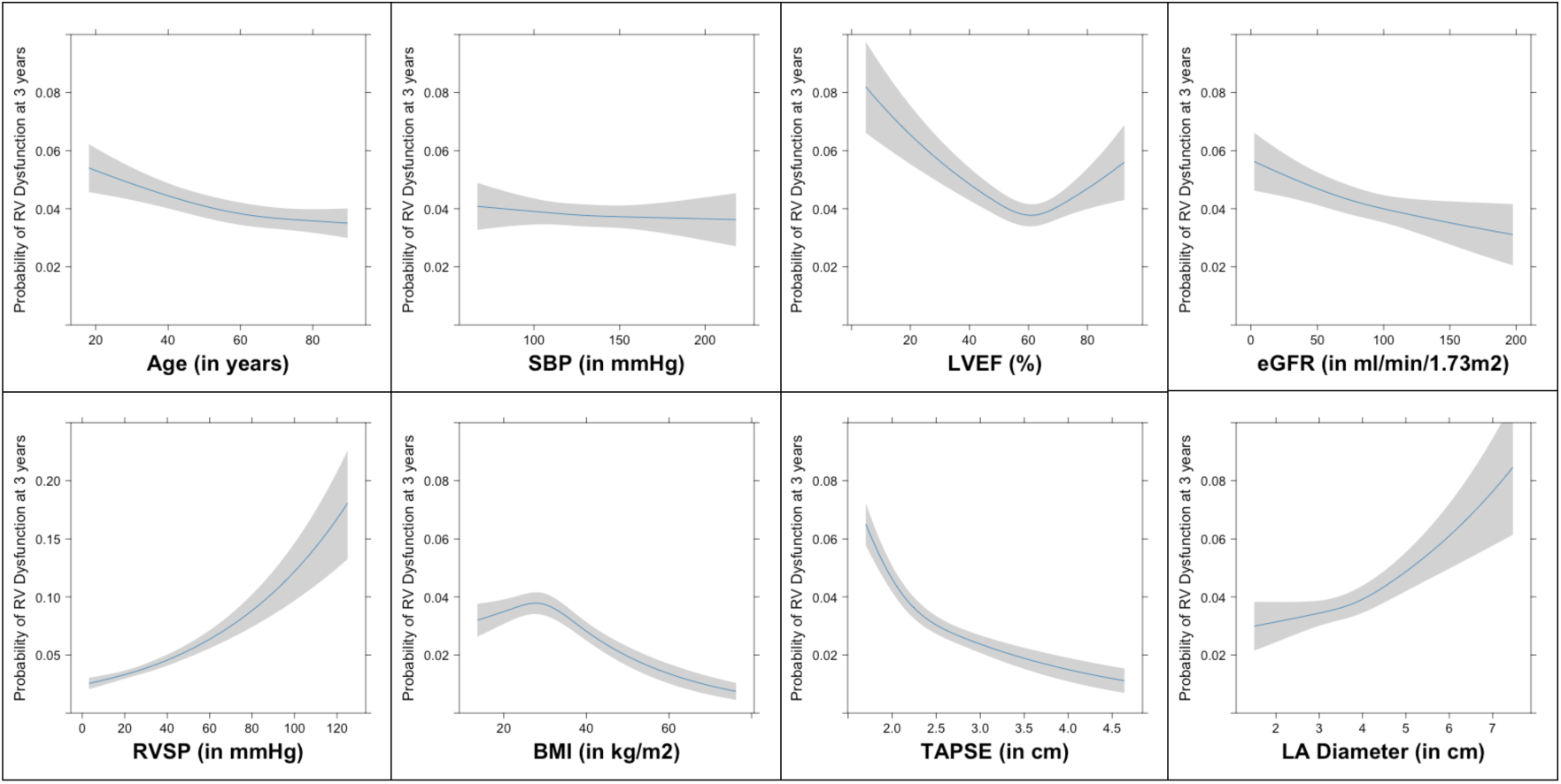
Associations between continuous variables at baseline and probability of right ventricular dysfunction at median follow up (3 years) Figure 3 displays the model-adjusted association between continuous baseline covariates and the outcome of incident right ventricular dysfunction as restricted cubic splines with 3 knots within the Full Cohort. The Y-axis displays the probability of normal RV dysfunction at 3 years of follow up and the x-axis displays the range of covariate values with the gray area representing a 95% confidence interval for the effect estimate at a given x-axis value.

On time-varying multivariable Cox regression, we found that the hazard of RVD associated with HF (HR 2.26 [95%CI 2.09–2.43) and AF (HR 1.81 [95%CI 1.69-2.94) become more pronounced **(eTable 2).** All baseline characteristics retained their directional association with hazard of incident RVD and all continuous variables retained their association with probability of RVD at 3 years in the Repeat TTE Cohort **(eTable 2, eFigure 1**).

We then assessed the association between baseline characteristics and incident RVD stratified by PH status **(eTable 3).** Several variables were more strongly associated with RVD in the PH-group, including male sex, Systemic Sclerosis, and left sided valvular disease. The relationships between continuous variables and probability of normal RV function (i.e shape of the splines) were similar between PH+ and PH-cohorts (**eFigure 2**).

### Association of Baseline Characteristics with Change in TAPSE

In the Repeat TTE Cohort, the mean change in TAPSE was −2.44mm +/− 5.78mm. Baseline AF (β = −1.22 mm, p<0.001), left-sided valvular disease (β = −1.18 mm, p=0.001), CAD (β = −0.93 mm, p<0.001), and HF (β = −0.73 mm, p=0.001) were strongly associated with larger decline in TAPSE. Adjusted β-coefficients with 95%CIs for all variables are displayed in **eTable 4.** Male sex, Diabetes, ILD, and obstructive sleep apnea were associated with decline in TAPSE to a lesser degree (all p<0.05). Increasing age (β = 0.09 mm per 10 years, p=0.005), SBP (β = 0.05 mm per 10mmHg, p=0.038), and LVEF (β = 0.27 mm per 10% increase, p<0.001) were associated with a smaller decline in TAPSE. Baseline RVSP, BMI, and baseline TAPSE were non-linearly associated with change in TAPSE (all p<0.05). For RVSP, the change in TAPSE was stable at −0.1 until an inflection point near 35mmHg, above which increasing RVSP is linearly associated with progressive decline in TAPSE. TAPSE change was stable for BMI <30, however as BMI increases above 30 there is an association with increasing improvement in TAPSE. The model-estimated TAPSE change by all continuous variables are displayed in **eFigure 3**.

### Association between Incident RV Dysfunction and HF Hospitalization

Of the Repeat TTE cohort, 13,142 patients (95.7%) had follow-up encounters at our institution after their second TTE and 2,343 of these patients (17.8%) had a subsequent HF hospitalization or rehospitalization. Median time between TTEs was 791 days (IQR 374-1510) and median follow up after second TTE was 227 days (IQR 42-434). Incident RVD (vs no development of RVD) was associated with an increased hazard of HF hospitalization (HR 2.02; 95%CI 1.85-2.21), exhibiting a hazard ratio of similar magnitude to that of prevalent HF (**eTable 5**). There was an increased hazard of HF hospitalization beginning near a 5mm decrease in TAPSE that steadily increased with greater TAPSE decline (**Figure 4A**). When restricted to outpatient follow up TTEs the hazard of HF hospitalization associated with incident RVD (HR 2.05; 95%CI 1.86-2.27) and relationship between TAPSE and HF hospitalization were unchanged (spline not shown).

**Figure 4:**
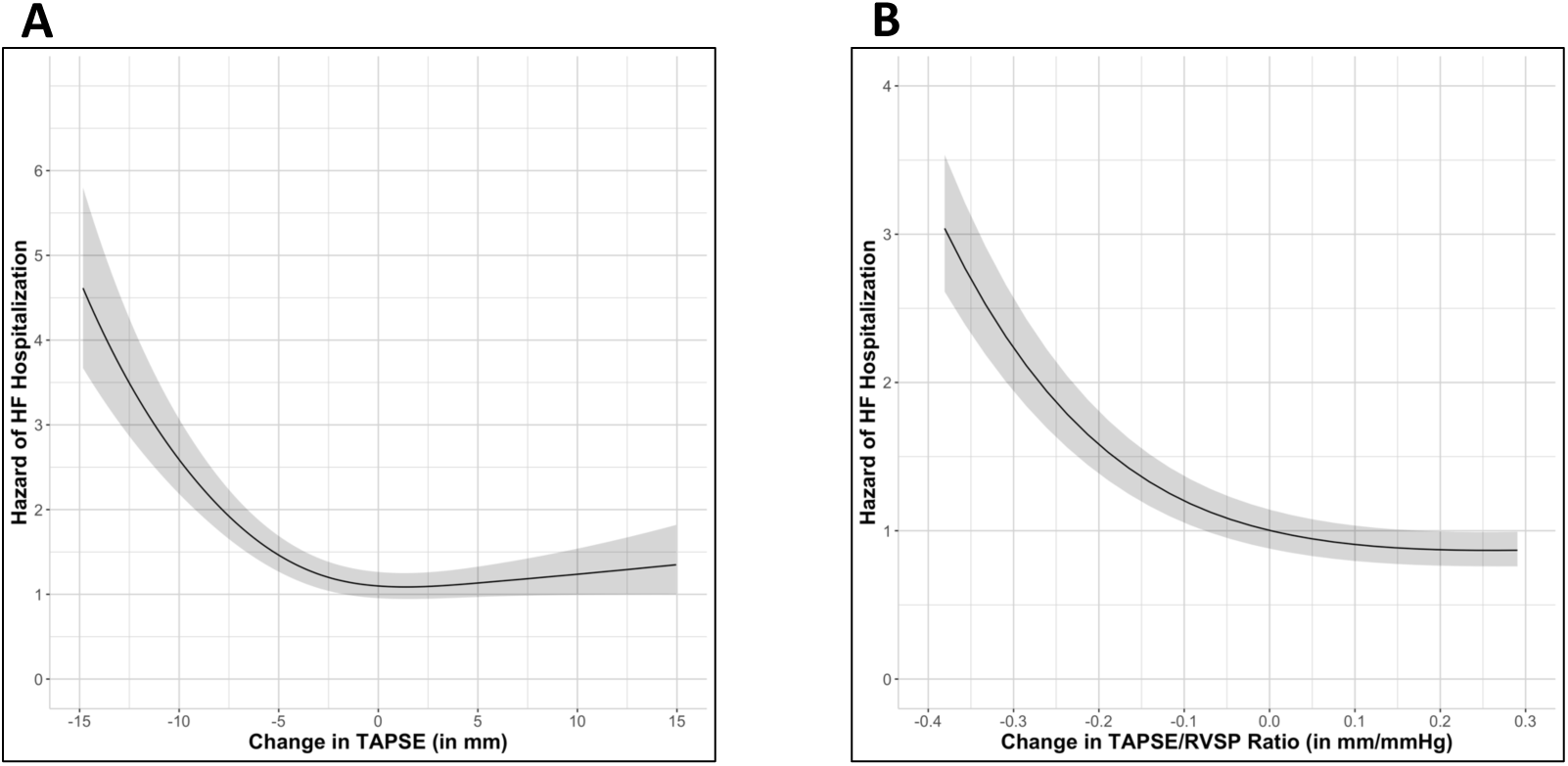
Association of change in (A) TAPSE and (B) TAPSE/RVSP Ratio with Hazard of HF Hospitalization/Rehospitalization. Figure 4 displays the Hazard of HF hospitalization or rehospitalization associated with a given change in (A) TAPSE or (B) TAPSE/RVSP Ratio, relative to the hazard of hospitalization or rehospitalization for no change.

### RV-PA Uncoupling

There were 44,563 patients from the full cohort with normal RV-PA coupling at baseline. Of these, 11,148 patients (25.0%) had an available follow up TTE with TASPE and RVSP measurement. New RV-PA uncoupling was uncovered in 1,592 patients (3.6%). The median time to development of RV-PA uncoupling was 869 days (IQR 381 – 1671), while patients who did not develop RV-PA uncoupling were followed for a median of 797 days (IQR 366 – 1572). The mean change in TAPSE/RVSP ratio was −0.11 mm/mmHg +/− 0.49.

The IR estimate of RV-PA uncoupling in the Full Cohort (minimum estimate) was 1.2 per 100 person-years (95%CI 1.1–1.3) and increased to 3.4 per 100 person-years (95%CI 3.2 – 3.6) in patients with a repeat RV-PA measurement. IRs of RV-PA uncoupling were substantially higher in the PH+ population and increased consistently as RVSP increased (**Table 2**). The baseline characteristics associated with RV-PA coupling were the same as the characteristics associated with RV dysfunction (**eTable 6, eFigure 4**). Degree of diastolic dysfunction, AF, HF, and systemic sclerosis demonstrated particularly strong associations with incident RV-PA uncoupling. As with RV dysfunction, we found that the hazard of RV-PA uncoupling associated with HF and AF become more pronounced on time-varying regression modeling. Incident RV-PA uncoupling was associated with an increased hazard of HF hospitalization (HR 3.21; 95%CI 2.84-3.63), exhibiting a hazard ratio greater than that of prevalent HF (**eTable 7**). There was an increased hazard of HF hospitalization beginning near a 0.1mm/mmHg decrease in TAPSE/RVSP ratio that steadily increased with further decline (**Figure 4B**).

## Discussion

We examined longitudinal development of incident RVD defined by TAPSE and RV-PA uncoupling defined by TAPSE/RVSP ratio in a large echocardiographic referral cohort. This work is significant because incidence rates for RVD uncoupling have not been reported, in contrast to virtually all other cardiopulmonary conditions. The major findings are: 1) The incidence of RVD in the echocardiographic referral population is substantial and 1.7-2.1 fold higher in individuals with PH (5.6-11.9/100 PY) compared to those without (2.7-7.2/100PY). 2) Among clinical characteristics, cardiovascular comorbidities were the most strongly associated with increased hazard of RVD and RV-PA uncoupling, with a possible causal relationship for HF and AF. 3) There appears to be a threshold effect for the association between RVSP and RVD near 35 mmHg. 4) Incident RVD and RV-PA uncoupling are independently associated with a 2-fold and 3-fold increased hazard of HF hospitalization, respectively. 5) Decline in TAPSE greater than 5mm and a decline in TAPSE/RVSP greater than 0.1 mm/mmHg are associated with an increased hazard of HF hospitalization.

Despite the well-established prognostic significance of RVD, it is one of the few cardiovascular phenotypes lacking data on IRs. Population-level prevalence estimates of RVD derive from longitudinal community-based NIH cohorts. Among 1,004 ARIC participants in their 70s with moderate comorbidity burden the prevalence of RVD by TTE 3D RVEF or RV longitudinal strain was 17.2%-19.8%,^17^ but significantly lower at 8% when measured by TAPSE or RV S’ among 3,433 CARDIA participants with low comorbidity burden^18^ and as low as 5.9% in MESA when measured by CMR RVEF.^16^ While there is some data on RV-PA coupling variation in the general population from ARIC (measured as echocardiographic RVEF/PASP),^17^ prevalence estimates of RV-PA uncoupling are lacking, in part due to the lack of consensus cutoff. The prevalence of RVD is substantial in selected populations such as HFpEF (26-49%),^32^ HFrEF (35-50%),^31,33–36^, and PH (>25%)^37^, as is the prevalence of RV-PA uncoupling which varies widely from 14-40% in the largest published cohorts using a TAPSE/PASP definition of <0.36mm/mmHg.^9,38^ To the best of our knowledge, there are no studies that estimate IRs for RVD or RV-PA uncoupling as in our study.

We found the rate of RVD to be 3.2-8.2% per person-year in a large, diverse echocardiographic referral cohort, increasing to 9.9-17.3% per person-year in patients with RVSP>60 mmHg. RV-PA uncoupling rates were generally lower, estimated between 1.2-3.4% per person-year for the full cohort and similarly increasing rapidly with higher RVSP. The minimum estimates are conservative because they assume all patients who did not undergo a repeat TTE did not develop RVD, which is unlikely to be true. Conversely, the maximum estimates are likely inflated by selection bias by including only patients referred for a repeat TTE. The true incident rate likely falls somewhere within the range. Despite inherent enrichment for cardiac comorbidities and structural disease in a TTE referral cohort our findings are highly relevant for clinical care in patients with suspected cardiopulmonary disease with an indication for TTE. The IR estimates and adjusted hazards for RVD described in this study identify patients at risk and provide an evidence base for powering future trials targeting RV function.

We identified cardiovascular comorbidities and cardiac structural disease as key drivers of both RVD and RV-PA uncoupling, confirming previously noted strong associations with HF, LV diastolic and systolic dysfunction, AF, coronary artery disease, and left-sided valvular disease.^7,9,10,32,35,39–42^ Tricuspid regurgitation was associated with decreased RVD, which may be related to preservation in lateral RV systolic motion via increased RV preload, RV dilation and increased stroke volume along with co-occurring compensatory RV hypertrophy. We found stronger associations between HF/AF and incident RVD when new cases were accounted for, suggesting a possible mechanistic link.^43^ Contrary to prior observations of a linkage between metabolic dysfunction and RVD, we found that higher BMI was associated with improved RV function. This relationship which may be specific to TAPSE however, as other RV function metrics decline with increasing BMI.^18,44–46^ Systemic sclerosis and left-sided valvular disease were associated with RVD in patients without PH, but not patients with PH, highlighting the need for future research.

We observed a threshold effect of RVSP on TAPSE near 35 mmHg, mirroring the relationship between RVSP and mortality,^5,47^ and correlating with the ERS/ERC 2022 guideline cutoff to suggest increased suspicion of PH at TRV >2.8 m/s (equivalent to RVSP of ∼34mmHg assuming a normal RAP).^24^ An RVSP ≥35 mmHg may represent a population-level pathologic threshold for RVD that warrants serial monitoring over time. Our finding lends further evidence to the appropriateness of the established cutoff for suspicion of PH.

Incident RVD and RV-PA uncoupling were strongly independently associated with increased HF hospitalization hazard after robust multivariable adjustment and on sensitivity analysis among patients with outpatient TTEs. Our results align with prior studies that have established prevalent RVD and RV-PA uncoupling as important prognostic findings in PH^5,48–51^ and HF.^9,31,52^ We define novel numeric cutoffs for HF hospitalization risk, noting an increased HF hazard with a TAPSE decline of 5mm or greater and TAPSE/RVSP decline of 0.1mm/mmHg or greater compared to no change over time.

### Limitations

Our cohort was derived from a TTE referral population, which introduces selection bias, but our findings are directly applicable to routine care. The IRs of RVD that we describe may be overestimated as not all patients with PH undergo TTE, potentially missing cases. Conversely, follow up TTEs were conducted according to clinical practice and RVD may have occurred before being identified, therefore incident rates may be systemically underestimated. Overall, these effects are unlikely to substantially change the estimated IRs. We chose TAPSE as the primary metric of RV function due to its widespread availability, reproducibility, and extensive prior study.^1,35,53^ Alternative TTE measures of RV function such as tricuspid annular velocity, fractional area change, or RV ejection fraction were less available in our cohort. We cannot determine whether beat-to-beat variability in TAPSE due to AF at the time of TTE affected our measures, but given the cohort size, this potential source of error likely did not impact our results. The associations observed in our study do not imply causality and should be considered hypothesis generating for future mechanistic studies. There is minimal data missingness in our primary analyses (<10% for all variables) that is unlikely to impact the relationships identified between baseline characteristics and incident RVD.

### Conclusions

We report the first incident rate estimates of RVD and RV-PA uncoupling among patients seeking care. The rate of incident RVD is substantial and may warrant closer monitoring among patients with RVSP >35mmHg. Cardiovascular comorbidities, especially HF and AF, increase the hazard of RVD and RV-PA uncoupling. RVD and RV-PA uncoupling are the strongest predictors of HF hospitalization even after accounting for prevalent HF and other cardiovascular comorbidities. A decline in TAPSE of greater than 5mm or a decline in TAPSE/RVSP of greater than 0.1 mm/mmHg warrant increased scrutiny and clinical surveillance.

## Data Availability

All data produced in the present study are available upon reasonable request to the authors

## Abbreviation List

AF: Atrial Fibrillation
BMI: Body Mass Index
CAD: Coronary Artery Disease
eGFR: Estimated Glomerular Filtration Rate
HF: Heart Failure
HFpEF: Heart Failure with preserved ejection fraction
HFrEF: Heart Failure with reduced ejection fraction
ILD: Interstitial Lung Disease
IR: Incidence Rate
LV: Left Ventricle
PH: Pulmonary Hypertension
RAP: Right atrial pressure
RV: Right Ventricle
RVD: Right Ventricular dysfunction
RV-PA: Right Ventricular – Pulmonary Artery
RVSP: Right Ventricular Systolic Pressure
TTE: Transthoracic Echocardiogram
TAPSE: Tricuspid Annular Plane Systolic Excursion
TRV: Tricuspid Regurgitant Velocity

