## Supplemental Materials for "Incidence of Right Ventricular Dysfunction in an Echocardiographic Referral Cohort"

*Table of Contents:*

Supplemental Methods: Page 2-3

eTable 1-7: Pages 4-10

eFigures 1-4: Pages 11-14

Supplemental References: Page 15

**Supplemental Methods**

*Exposures, Covariates, and Outcomes*

Normal RV function was defined by tricuspid annular plane systolic excursion (TAPSE) ≥17mm based on the 2015 ASE/EACI guidleines.[1] We selected TAPSE as measure of RV systolic function in this study rather than other TTE parameters such as fractional area change or tricuspid annular velocity (S') since these were less consistently measured in our clinical dataset and require high image quality. Last medical contact was defined as last date for any measurement, procedure, condition, or visit.

From each TTE we abstracted TAPSE, TRV, RAP, PASP, left atrial diameter, average E/e', diastolic function, significant valvular disease, and left ventricular ejection fraction (LVEF). We selected body mass index (BMI), systolic blood pressure (SBP), and estimated glomerular filtration rate (eGFR) measurements within 180-days before or after baseline TTE. We extracted data on comorbidities implicated in the development of chronic RVD, including heart failure (HF), coronary artery disease (CAD), atrial fibrillation (AF), interstitial lung disease (ILD), chronic obstructive pulmonary disease (COPD), obstructive sleep apnea (OSA), human immunodeficiency virus (HIV), diabetes mellitus (DM), hyperlipidemia (HLD), and systemic sclerosis. Comorbidities were considered present at baseline If a patient had one inpatient or two outpatient ICD-9 or 10 codes dated before the baseline TTE or within 180 days afterwards. We categorized valvular disease dichotomously, as present or absent based on moderate or worse left-sided valvular disease and tricuspid regurgitation (TR), with mild mitral stenosis also included as significant left-sided valvular disease. PASP was calculated from TRV and right atrial pressure (RAP) when unavailable from the TTE report using the modified Bernoulli equation. We assumed a RAP of 5 mmHg when missing, based on prior evidence.[2,3]

*Characteristics Associated with RVD and TAPSE*

For the Full Cohort, the outcome measure was dichotomous presence or absence of RVD. The duration of exposure was the time from baseline TTE to follow up TTE with RVD, or for those without development of RVD the time from baseline TTE to last medical contact. For the Repeat TTE cohort we selected the TAPSE value from the temporally closest TTE (to baseline TTE) with RVD for patients who developed RVD and the TAPSE from the latest TTE available for patients who did not develop RVD, excluding patients without an available follow up TAPSE. Data regarding renal function and echocardiographic surrogates of diastolic dysfunction had a high degree of missingness therefore were not included in the primary models, but were individually added to the primary model in sensitivity analyses.

*HF Hospitalization*

We defined HF hospitalization by inpatient ICD-9 codes 425 or 428 or ICD-10 codes I42 or I50. The time of the second TTE was considered baseline. If the TTE was conducted inpatient, the next hospitalization for HF was considered as the outcome event. Inpatient TTE status determined the presence of any inpatient ICD code within one day of the TTE. Patients without hospitalization were censored at the time of last medical touch.

| eTable 1: Clinical Characteristics at Baseline Transthoracic Echocardiogram by Follow Up Status | | | |
|---|---|---|---|
| | **No Follow Up Echo (n = 32018)** | **Follow Up Echo (n = 13735)** | **p-value** |
| **Age in years** | 62 (49–72) | 64 (51–72) | <0.001 |
| **Male Sex** | 13994 (44%) | 6725 (49%) | <0.001 |
| **BMI in kg/m$^2$** | 28.7 (24.8–33.8) | 29.0 (25.1–33.9) | <0.001 |
| **Race/Ethnicity** | | | <0.001 |
| White, Non-Hispanic | 25538 (80%) | 11137 (81%) | |
| Black | 3883 (12%) | 1881 (14%) | |
| Hispanic | 459 (1%) | 191 (1%) | |
| Other | 1145 (4%) | 407 (3%) | |
| **Baseline TAPSE in cm** | 2.20 (1.99–2.51) | 2.20 (1.93–2.50) | <0.001 |
| **Baseline RVSP in mmHg** | 28 (23–35) | 30 (24–37) | <0.001 |
| **eGFR in mL/min/1.73m$^2$** | 79 (61–95) | 75 (55–92) | <0.001 |
| **SBP in mmHg** | 128 (116–142) | 126 (114–140) | <0.001 |
| **Systemic Sclerosis** | 168 (1%) | 158 (1%) | <0.001 |
| **Heart Failure** | 6730 (21%) | 5459 (40%) | <0.001 |
| **LV Ejection Fraction** | 62 (57–67) | 60 (55–66) | <0.001 |
| **Atrial fibrillation** | 6919 (22%) | 4171 (30%) | <0.001 |
| **Coronary Artery Disease** | 8411 (26%) | 5183 (38%) | <0.001 |
| **Left-Sided Valvular Disease** | 796 (2%) | 1301 (9%) | <0.001 |
| **Tricuspid Regurgitation** | 431 (1%) | 432 (3%) | <0.001 |
| **Interstitial Lung Disease** | 3809 (12%) | 2526 (18%) | <0.001 |
| **COPD** | 3888 (12%) | 1901 (14%) | <0.001 |
| **Obstructive Sleep Apnea** | 5900 (18%) | 2989 (22%) | <0.001 |
| **HIV** | 342 (1%) | 158 (1%) | <0.001 |
| **Diabetes Mellitus** | 7827 (24%) | 4125 (30%) | <0.001 |
| **Hyperlipidemia** | 17519 (55%) | 8448 (62%) | <0.001 |
| **Follow Up Time in days** | 623 (297–1267) | 1137 (570–1990) | <0.001 |

**eTable 2: Multivariate Cox Regression Model of Characteristics associated with Incident Right Ventricular Dysfunction**

| | Full Cohort | Full Cohort Time-Updated[c] | Repeat TTE Cohort | Repeat TTE Cohort Time-Updated[c] |
|---|---|---|---|---|
| | Hazard Ratio [95% CI] | | | |
| **Age[a]** | 0.89 (0.84–0.94)[b] | 0.76 (0.69–0.85)[b] | 1.00 (0.94–1.06)[b] | 0.96 (0.90–1.01)[b] |
| **Male Sex** | 1.09 (1.02–1.16) | 0.94 (0.84–1.05) | 1.07 (1.00–1.14) | 1.03 (0.97–1.10) |
| **BMI[a]** | 0.93 (0.88–0.98)[b] | 0.88 (0.83–0.93)[b] | 0.90 (0.85–0.95)[b] | 0.87 (0.82–0.91)[b] |
| **Race/Ethnicity (vs. Non-Hispanic White)** | | | | |
| Black | 0.93 (0.84–1.02) | 0.92 (0.84–1.02) | 0.88 (0.80–0.97) | 0.88 (0.80–0.97) |
| Hispanic | 1.03 (0.79–1.35) | 1.05 (0.80–1.38) | 1.00 (0.77–1.32) | 1.23 (0.78–1.34) |
| Other | 0.99 (0.82–1.19) | 1.01 (0.84–1.22) | 1.03 (0.85–1.23) | 1.05 (0.87–1.27) |
| **Baseline TAPSE[a]** | 0.64 (0.61–0.67)[b] | 0.64 (0.61–0.67)[b] | 0.60 (0.57–0.64)[b] | 0.62 (0.59–0.65)[b] |
| **Baseline RVSP[a]** | 1.22 (1.17–1.28)[b] | 1.18 (1.06–1.32)[b] | 1.13 (1.08–1.19)[b] | 1.12 (1.06–1.17)[b] |
| **Systolic Blood Pressure[a]** | 0.98 (0.94–1.02) | 0.94 (0.87–1.01) | 0.99 (0.95–1.03) | 1.00 (0.95–1.04) |
| **Systemic Sclerosis** | 1.84 (1.41–2.39) | 1.81 (1.41–2.32) | 1.31 (1.00–1.71) | 1.32 (1.03–1.70) |
| **Heart Failure** | 1.88 (1.75–2.03) | 2.26 (2.09–2.43) | 1.43 (1.33–1.53) | 1.59 (1.48–1.71) |
| **LV Ejection Fraction[a]** | 1.01 (0.96–1.06)[b] | 1.02 (0.98-1.07)[b] | 1.07 (1.02-1.12)[b] | 1.08 (1.03-1.12)[b] |
| **Atrial fibrillation** | 1.54 (1.44–1.65) | 1.81 (1.69–2.94) | 1.44 (1.34–1.53) | 1.61 (1.50–1.72) |
| **Coronary Artery Disease** | 1.26 (1.17–1.36) | 1.39 (1.29–1.50) | 1.18 (1.10–1.27) | 1.28 (1.19–1.38) |
| **Left-Sided Valvular Disease** | 1.68 (1.53–1.85) | 1.58 (1.44–1.74) | 1.26 (1.15–1.38) | 1.23 (1.12–1.35) |
| **Tricuspid Regurgitation** | 0.84 (0.72–0.98) | 0.90 (0.75–1.07) | 0.82 (0.71–0.96) | 0.82 (0.70–0.96) |
| **Interstitial Lung Disease** | 1.41 (1.30–1.52) | 1.53 (1.42–1.64) | 1.27 (1.17–1.37) | 1.36 (1.26–1.46) |
| **Chronic Obstructive Lung Disease** | 1.07 (0.98–1.17) | 1.02 (0.94–1.11) | 1.14 (1.04–1.24) | 1.07 (0.99–1.16) |
| **Obstructive Sleep Apnea** | 1.09 (1.00–1.18) | 1.09 (0.81–1.39) | 1.02 (0.94–1.10) | 1.01 (0.94–1.09) |
| **HIV** | 1.12 (0.85–1.46) | 1.06 (0.81–1.39) | 1.16 (0.89–1.53) | 1.13 (0.87–1.49) |
| **Diabetes Mellitus** | 1.30 (1.21–1.39) | 1.29 (1.21–1.38) | 1.17 (1.18–1.37) | 1.27 (1.19–1.37) |
| **Hyperlipidemia** | 1.05 (0.97–1.13) | 1.07 (0.98–1.16) | 1.00 (0.92–1.08) | 0.98 (0.90–1.06) |
| **Sensitivity Analyses with >10% Missingness[d]** | | | | |
| **eGFR[a]** | 0.89 (0.85–0.94)[b] | 0.91 (0.87-0.96)[b] | 0.88 (0.84–0.92)[b] | 0.90 (0.86-0.95)[b] |
| **LA diameter[a]** | 1.16 (1.09–1.24)[b] | 1.11 (1.05-1.18) | 1.11 (1.04–1.18)[b] | 1.07 (0.94-1.10)[b] |
| **Diastolic Dysfunction (vs. normal)** | | | | |
| Stage 1 | 1.14 (1.01-1.29) | 1.07 (0.95-1.21) | 1.01 (0.90-1.14) | 1.02 (0.91-1.15) |
| Stage 2 | 1.30 (1.10-1.54) | 1.16 (0.98-1.38) | 1.22 (1.03-1.44) | 1.17 (1.01-1.36) |
| Stage 3 | 1.37 (1.07-1.75) | 1.26 (0.99-1.60) | 1.40 (1.10-1.78) | 1.36 (1.09-1.70) |
| **LV average E/e'[a]** | 1.02 (0.93-1.12) | 1.00 (0.92-1.10) | 0.97 (0.89–1.07) | 0.97 (0.89-1.06) |

[a]Hazard Ratios for continuous variables are displayed as 75th %ile compared to 25th %ile. Hazards may not reflect statistically significant association.
[b]Denotes a statistically significant association between continuous variable and outcome ($p < 0.05$)
[c]ICD-code derived comorbidities were time-updated in this model, including Heart Failure, Atrial Fibrillation, Coronary Artery Disease, Interstitial Lung Disease, Systemic Sclerosis, Chronic Obstructive Lung Disease, Obstructive Sleep Apnea, HIV, Diabetes Mellitus, and Hyperlipidemia
[d]Each variable below this sub header was added independently into the primary model of variables above

| eTable 3: Multivariate Cox Regression Model of Baseline Characteristics associated with Incident Right Ventricular Dysfunction, Stratified by PH Status | | | | |
|---|---|---|---|---|
| | Full Cohort | | Repeat TTE Cohort | |
| | PH+ | PH- | PH+ | PH- |
| **Baseline TAPSE[a]** | 0.65 (0.59–0.71)[b] | 0.63 (0.59–0.66)[b] | 0.66 (0.59–0.73)[b] | 0.59 (0.56–0.63)[b] |
| **Baseline RVSP[a]** | 1.20 (1.08–1.35)[b] | 1.11 (1.05–1.17)[b] | 1.21 (1.08–1.36)[b] | 1.05 (0.99–1.10) |
| **Age[a]** | 0.81 (0.72–0.90)[b] | 0.91 (0.85–0.98)[b] | 0.98 (0.88–1.09) | 0.98 (0.92–1.06)[b] |
| **Male Sex** | 0.98 (0.88–1.10) | 1.12 (1.03–1.21) | 0.99 (0.89–1.12) | 1.10 (1.02–1.19) |
| **BMI[a]** | 0.85 (0.78–0.93)[b] | 0.95 (0.89–1.02)[b] | 0.80 (0.74–0.88)[b] | 0.94 (0.88–1.00)[b] |
| **Race/Ethnicity (vs. Non–Hispanic White)** | | | | |
| Black | 0.93 (0.80–1.08) | 0.92 (0.81–1.04) | 0.89 (0.77–1.04) | 0.88 (0.78–0.99) |
| Hispanic | 1.34 (0.86–2.10) | 0.91 (0.65–1.28) | 1.39 (0.83–2.02) | 0.89 (0.63–1.25) |
| Other | 1.04 (0.74–1.45) | 0.99 (0.79–1.24) | 0.94 (0.67–1.31) | 1.09 (0.87–1.36) |
| **Systolic Blood Pressure[a]** | 0.92 (0.85–0.99) | 1.00 (0.96–1.06) | 0.94 (0.87–1.02) | 1.01 (0.96–1.07)[b] |
| **Diabetes Mellitus** | 1.15 (1.02–1.30) | 1.36 (1.25–1.49) | 1.18 (1.04–1.34) | 1.30 (1.19–1.42) |
| **Hyperlipidemia** | 0.97 (0.85–1.11) | 1.09 (0.99–1.19) | 0.93 (0.81–1.06) | 1.03 (0.94–1.14) |
| **Interstitial Lung Disease** | 1.25 (1.10–1.42) | 1.45 (1.32–1.60) | 1.18 (1.04–1.34) | 1.30 (1.18–1.43) |
| **Chronic Obstructive Lung Disease** | 1.00 (0.87–1.15) | 1.15 (1.02–1.29) | 1.06 (0.92–1.21) | 1.19 (1.06–1.34) |
| **Obstructive Sleep Apnea** | 1.14 (1.00–1.31) | 1.05 (0.95–1.16) | 1.05 (0.91–1.20) | 1.00 (0.91–1.11) |
| **Systemic Sclerosis** | 1.40 (0.94–2.10) | 2.17 (1.53–3.09) | 1.09 (0.73–1.63) | 1.45 (1.02–2.07) |
| **HIV** | 0.80 (0.45–1.42) | 1.23 (0.90–1.67) | 0.83 (0.47–1.48) | 1.28 (0.95–1.76) |
| **Heart Failure** | 1.74 (1.52–1.98) | 1.91 (1.74–2.09) | 1.52 (1.33–1.73) | 1.39 (1.27–1.51) |
| **Atrial fibrillation** | 1.62 (1.43–1.82) | 1.52 (1.40–1.66) | 1.55 (1.38–1.75) | 1.39 (1.28–1.51) |
| **Coronary Artery Disease** | 1.25 (1.10–1.42) | 1.26 (1.16–1.38) | 1.14 (1.01–1.30) | 1.18 (1.08–1.29) |
| **Left–Sided Valvular Disease** | 1.16 (1.00–1.35) | 2.25 (2.00–2.54) | 1.03 (0.88–1.20) | 1.43 (1.27–1.60) |
| **Tricuspid Regurgitation** | 0.85 (0.71–1.02) | 1.11 (0.83–1.47) | 0.81 (0.67–0.97) | 0.97 (0.73–1.29) |
| **LV Ejection Fraction[a]** | 0.94 (0.86–1.02)[b] | 1.03 (0.98–1.09)[b] | 0.99 (0.91–1.08)[b] | 1.10 (1.04–1.15)[b] |

[a]Hazard Ratios for continuous variables are displayed as 75th %ile compared to 25th %ile. Hazards may not reflect statistically significant association.
[b]Denotes a statistically significant association between a continuous variable (linear or non-linear) and the outcome ($p < 0.05$)

| eTable 4: Ordinary Least Square Regression Model of Baseline Characteristics associated with Change in TAPSE (in mm) | | |
|---|---|---|
| | Coefficient (S.E.) | p-value |
| **Baseline TAPSE** | Non-linear | <0.001 |
| **Baseline RVSP** | Non-linear | 0.016 |
| **Age (per 10 years)** | 0.09 (0.03) | 0.005 |
| **Male Sex** | 0.53 (0.18) | <0.001 |
| **BMI** | Non-linear | <0.001 |
| **Race/Ethnicity (vs. Non-Hispanic White)** | | |
| Black | 0.30 (0.13) | 0.021 |
| Hispanic | -0.77 (0.78) | 0.325 |
| Other | 0.39 (0.50) | 0.439 |
| **Systolic Blood Pressure (per 10 mmHg)** | 0.05 (0.02) | 0.038 |
| **Diabetes Mellitus** | - 0.41 (0.10) | <0.001 |
| **Hyperlipidemia** | 0.11 (0.11) | 0.283 |
| **Interstitial Lung Disease** | - 0.46 (0.12) | <0.001 |
| **Chronic Obstructive Lung Disease** | -0.10 (0.13) | 0.447 |
| **Obstructive Sleep Apnea** | 0.26 (0.11) | 0.022 |
| **Systemic Sclerosis** | -0.76 (0.04) | 0.060 |
| **HIV** | -0.42 (0.40) | 0.302 |
| **Heart Failure** | - 0.73 (0.10) | <0.001 |
| **Atrial fibrillation** | - 1.22 (0.10) | <0.001 |
| **Coronary Artery Disease** | - 0.93 (0.10) | <0.001 |
| **Left-Sided Valvular Disease** | - 1.18 (0.15) | <0.001 |
| **Tricuspid Regurgitation** | - 0.37 (0.26) | 0.200 |
| **LV Ejection Fraction (per 10% increment)** | 0.27 (0.04) | <0.001 |
| **Follow up (per year)** | -0.03 (0.02) | 0.087 |

| eTable 5: Multivariable Cox Regression of Characteristics Associated with HF Hospitalization (RV Dysfunction) | | |
|---|---|---|
| | Hazard Ratio | 95% CI |
| **Incident RV Dysfunction** | 2.02 | 1.84 – 2.21 |
| **Heart Failure**[b] | 2.00 | 1.81 – 2.22 |
| **Atrial Fibrillation**[b] | 1.20 | 1.10 – 1.32 |
| **Coronary Artery Disease**[b] | 1.29 | 1.17 – 1.42 |
| **Systemic Sclerosis**[b] | 1.25 | 0.88 – 1.78 |
| **Interstitial Lung Disease**[b] | 1.17 | 1.05 – 1.29 |
| **Chronic Obstructive Pulmonary Disease**[b] | 1.41 | 1.27 – 1.57 |
| **Obstructive Sleep Apnea**[b] | 1.11 | 1.00 – 1.22 |
| **Human Immunodeficiency Virus**[b] | 1.35 | 0.98 – 1.85 |
| **Diabetes Mellitus**[b] | 1.49 | 1.36 – 1.64 |
| **Hyperlipidemia**[b] | 0.89 | 0.80 – 0.99 |
| **Baseline Tricuspid Regurgitation**[b] | 0.74 | 0.61 – 0.91 |
| **Baseline Left Sided Valvular Disease**[b] | 1.03 | 0.91 – 1.17 |
| **Age at 2nd TTE (per 10 years)** | 1.03 | 0.96 – 1.10 |
| **Male Sex** | 1.04 | 0.95 – 1.13 |
| **Baseline BMI**[a] | 1.06 | 0.99 – 1.13 |
| **Race (vs White Race)** | | |
| Black | 1.22 | 1.09 – 1.35 |
| Hispanic | 1.24 | 0.86 – 1.78 |
| Other | 0.96 | 0.73 – 1.26 |
| **Baseline TAPSE (per 10 mm)** | 1.31 | 1.01 – 1.70 |
| **Baseline Systolic Blood Pressure (per 10 mmHg)** | 0.99 | 0.94 – 1.04 |
| **Baseline RVSP (per 10 mmHg)** | 1.30 | 1.21 – 1.40 |
| **Baseline LVEF (per 10%)** | 0.88 | 0.83 – 0.92 |

[a]Hazard Ratios for continuous variables found to be non-linearly associated with the outcome are displayed as 75th %ile compared to 25th %ile. Hazards may not reflect statistically significant association.
[c]Comorbidities are included as time-varying covariates in the model

| eTable 6: Multivariate Cox Regression Model of Characteristics associated with RV-PA Uncoupling | | |
|---|---|---|
| | **Full Cohort** | **Full Cohort Time-Updated**[c] |
| | **Hazard Ratio [95% CI]** | |
| **Age**[a] | 1.08 (0.97 – 1.20)[b] | 0.98 (0.89 – 1.08)[b] |
| **Male Sex** | 1.11 (0.99 – 1.25) | 1.04 (0.92 – 1.17) |
| **BMI**[a] | 0.99 (0.90 – 1.10)[b] | 0.91 (0.83 – 1.01)[b] |
| **Race/Ethnicity (vs. Non-Hispanic White)** | | |
| Black | 1.18 (1.01 – 1.39) | 1.18 (1.00 – 1.39) |
| Hispanic | 0.90 (0.52 – 1.56) | 0.91 (0.53 – 1.59) |
| Other | 0.95 (0.64 – 1.41) | 0.94 (0.63 – 1.39) |
| **Baseline TAPSE**[a] | 0.64 (0.59 – 0.70)[b] | 0.66 (0.61 – 0.72)[b] |
| **Baseline RVSP**[a] | 2.43 (2.14 – 2.75)[b] | 2.23 (1.97 – 2.52)[b] |
| **Systolic Blood Pressure**[a] | 0.96 (0.89 – 1.04) | 0.98 (0.91 – 1.06) |
| **Systemic Sclerosis** | 2.67 (1.71 – 4.18) | 2.45 (1.61 – 3.72) |
| **Heart Failure** | 1.94 (1.69 – 2.23) | 3.05 (2.63 – 3.54) |
| **LV Ejection Fraction**[a] | 0.90 (0.92 – 0.99)[b] | 0.93 (0.84 – 1.02)[b] |
| **Atrial fibrillation** | 1.76 (1.55 – 1.99) | 2.22 (1.95 – 2.52) |
| **Coronary Artery Disease** | 1.29 (1.13 – 1.48) | 1.40 (1.23 – 1.61) |
| **Left-Sided Valvular Disease** | 1.55 (1.33 – 1.81) | 1.42 (1.22 – 1.66) |
| **Tricuspid Regurgitation** | 1.02 (0.81 – 1.27) | 1.04 (0.83 – 1.31) |
| **Interstitial Lung Disease** | 1.09 (0.95 – 1.26) | 1.33 (1.17 – 1.51) |
| **Chronic Obstructive Lung Disease** | 1.26 (1.08 – 1.46) | 1.26 (1.10 – 1.44) |
| **Obstructive Sleep Apnea** | 0.99 (0.85 – 1.15) | 1.07 (0.93 – 1.22) |
| **HIV** | 0.98 (0.58 – 1.68) | 0.89 (0.52 – 1.51) |
| **Diabetes Mellitus** | 1.37 (1.21 – 1.56) | 1.39 (1.22 – 1.57) |
| **Hyperlipidemia** | 0.90 (0.78 – 1.04) | 0.85 (0.73 – 1.00) |
| **Sensitivity Analyses with >10% Missingness**[d] | | |
| **eGFR**[a] | 0.82 (0.75 – 0.89)[b] | 0.84 (0.77 – 0.91)[b] |
| **LA diameter**[a] | 1.21 (1.07 – 1.36)[b] | 1.13 (1.00 – 1.28)[b] |
| **Diastolic Dysfunction (vs. normal)** | | |
| Stage 1 | 1.32 (1.06 – 1.65) | 1.19 (0.95 – 1.48) |
| Stage 2 | 1.92 (1.47 – 2.51) | 1.52 (1.17 – 1.98) |
| Stage 3 | 2.05 (1.42 – 3.00) | 1.66 (1.16 – 2.38) |
| **LV average E/e'**[a] | 1.18 (0.97 – 1.44) | 1.14 (0.95 – 1.38) |

[a]Hazard Ratios for continuous variables are displayed as 75[th] %ile compared to 25[th] %ile. Hazards may not reflect statistically significant association.
[b]Denotes a statistically significant association between continuous variable and outcome (p <0.05)
[c]ICD-code derived comorbidities were time-updated in this model, including Heart Failure, Atrial Fibrillation, Coronary Artery Disease, Interstitial Lung Disease, Systemic Sclerosis, Chronic Obstructive Lung Disease, Obstructive Sleep Apnea, HIV, Diabetes Mellitus, and Hyperlipidemia
[d]Each variable below this sub header was added independently into the primary model of variables above

| eTable 7: Multivariable Cox Regression of Characteristics Associated with HF Hospitalization (RV-PA Uncoupling) | | |
|---|---|---|
| | Hazard Ratio | 95% CI |
| **RV-PA Uncoupling** | 3.21 | 2.84 – 3.63 |
| **Heart Failure**[b] | 1.95 | 1.71 = 2.22 |
| **Atrial Fibrillation**[b] | 1.11 | 0.98 – 1.25 |
| **Coronary Artery Disease**[b] | 1.28 | 1.13 – 1.46 |
| **Systemic Sclerosis**[b] | 1.42 | 0.89 – 2.28 |
| **Interstitial Lung Disease**[b] | 1.37 | 1.20 – 1.56 |
| **Chronic Obstructive Pulmonary Disease**[b] | 1.22 | 1.06 – 1.41 |
| **Obstructive Sleep Apnea**[b] | 1.17 | 1.02 – 1.33 |
| **Human Immunodeficiency Virus**[b] | 1.22 | 0.82 – 1.81 |
| **Diabetes Mellitus**[b] | 1.51 | 1.34 – 1.70 |
| **Hyperlipidemia**[b] | 0.98 | 0.86 – 1.14 |
| **Baseline Tricuspid Regurgitation**[b] | 0.81 | 0.62 – 1.05 |
| **Baseline Left Sided Valvular Disease**[b] | 1.09 | 0.94 – 1.26 |
| **Age at 2$^{nd}$ TTE (per 10 years)** | 1.06 | 0.97 – 1.16 |
| **Male Sex** | 1.07 | 0.95 – 1.20 |
| **Baseline BMI**[a] | 0.98 | 0.96 – 1.00 |
| **Race (vs White Race)** | | |
| Black | 1.19 | 1.03 – 1.38 |
| Hispanic | 1.05 | 0.65 – 1.71 |
| Other | 0.72 | 0.48 – 1.10 |
| **Baseline TAPSE (per 10 mm)** | 1.30 | 0.92 – 1.83 |
| **Baseline Systolic Blood Pressure (per 10 mmHg)** | 1.02 | 0.93 – 1.11 |
| **Baseline RVSP (per 10 mmHg)** | 1.25 | 1.09 – 1.44 |
| **Baseline LVEF (per 10%)** | 0.90 | 0.85 – 0.96 |

[a]Hazard Ratios for continuous variables found to be non-linearly associated with the outcome are displayed as 75$^{th}$ %ile compared to 25$^{th}$ %ile. Hazards may not reflect statistically significant association.
[c]Comorbidities are included as time-varying covariates in the model

**eFigure 1**: Associations between continuous variables and the probability of right ventricular dysfunction at 3 years in the Repeat TTE Cohort

eFigure 1 displays the model-adjusted association between continuous baseline covariates and the outcome of incident right ventricular dysfunction as restricted cubic splines with 3 knots within the Repeat TTE Cohort. The Y-axis displays the probability of RV dysfunction at 3 years of follow up and the x-axis displays the range of covariate values with the gray area representing a 95% confidence interval for the effect estimate at a given x-axis value. Relationships between continuous covariates and probability of right ventricular dysfunction (the shape of the curves) in the Repeat TTE Cohort are essentially unchanged from the full cohort (Figure 3).

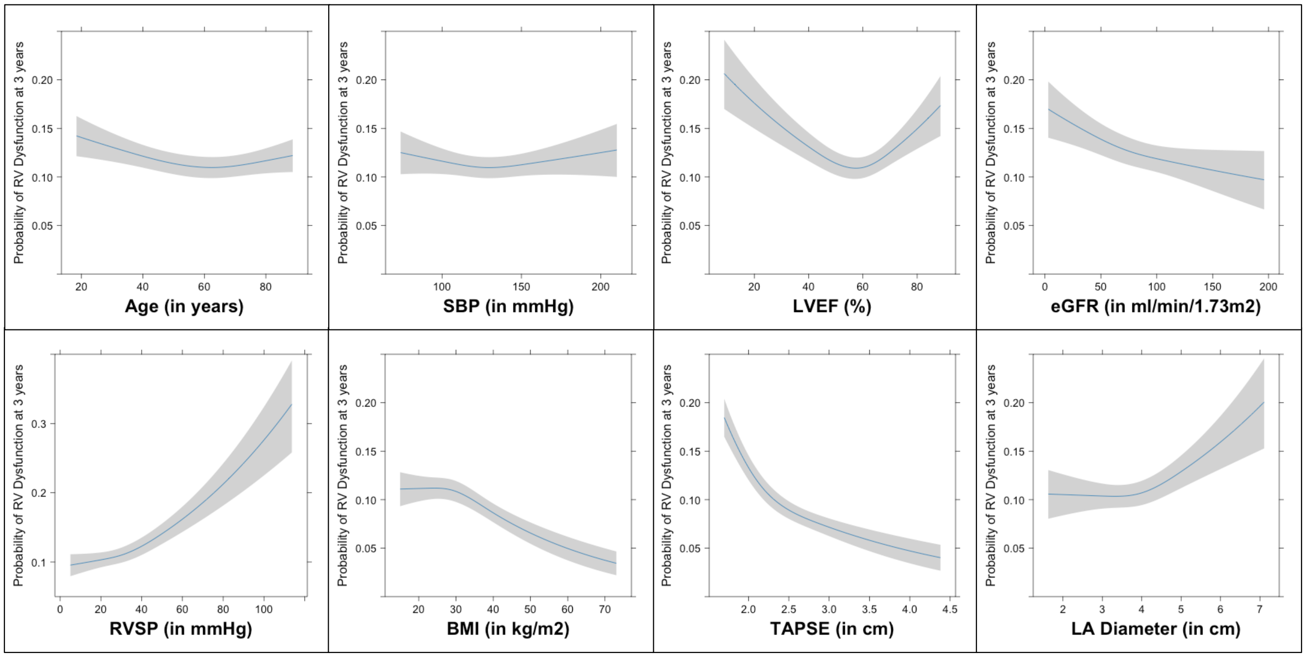

**eFigure 2:** Associations between continuous variables and the probability of right ventricular dysfunction at 3 years, comparing significantly associated variables between PH+ and PH- cohorts.

eFigure 2 displays the model-adjusted association between continuous baseline covariates and the outcome of incident right ventricular dysfunction as restricted cubic splines with 3 knots. Only those covariates with a statistically significant association with right ventricular dysfunction are displayed. The Y-axis displays the probability of right ventricular dysfunction at 3 years of follow up and the x-axis displays the range of covariate values with the gray area representing a 95% confidence interval for the effect estimate at a given x-axis value. Relationships between continuous covariates and probability of right ventricular dysfunction (the shape of the curves) are similar between the PH+ and PH- cohorts.

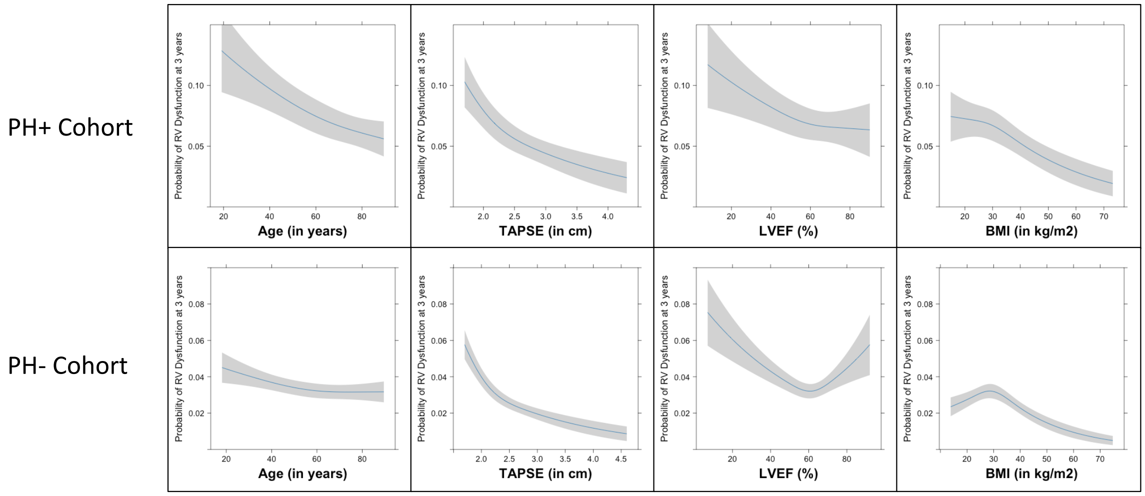

### **eFigure 3:** Associations between continuous variables and change in TAPSE

eFigure 3 displays the model-adjusted association between continuous baseline covariates and the outcome of change in TAPSE as restricted cubic splines with 3 knots. The Y-axis displays the change in TAPSE and the x-axis displays the range of covariate values with the gray area representing a 95% confidence interval for the effect estimate at a given x-axis value.

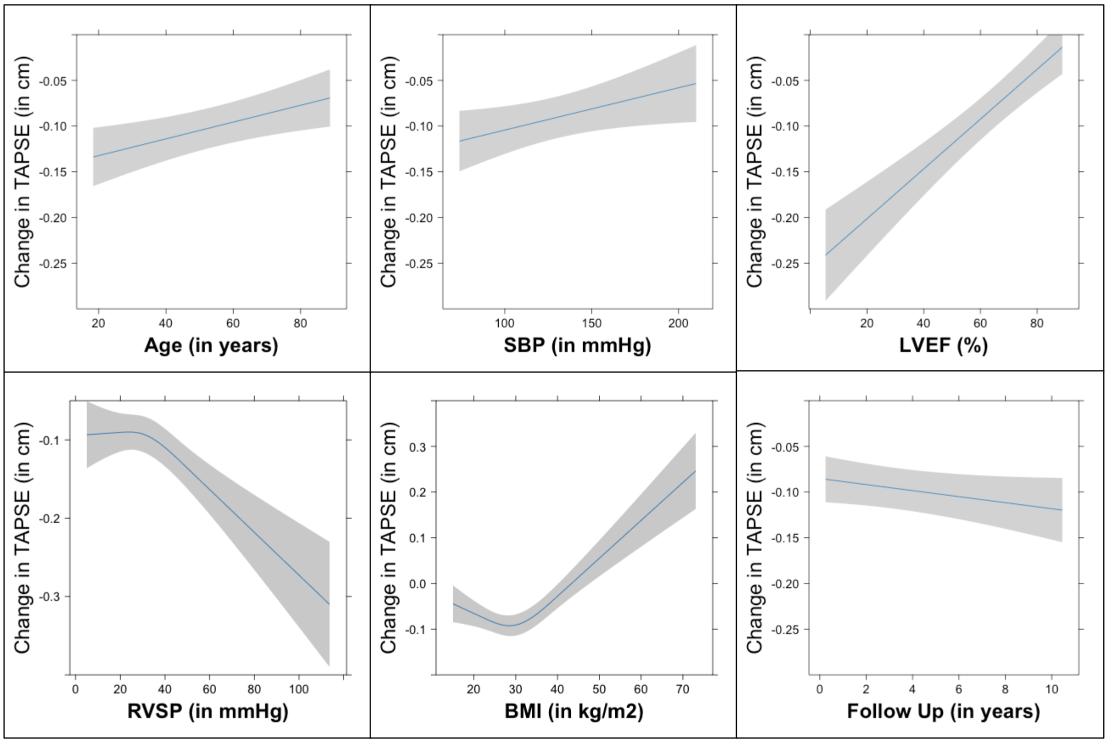

**eFigure 4**: Associations between continuous variables and the probability of RV-PA Uncoupling at 3 years in the full cohort

eFigure 4 displays the model-adjusted association between continuous baseline covariates and the outcome of RV-PA uncoupling as restricted cubic splines with 3 knots within the Full Cohort. The Y-axis displays the probability of RV-PA uncoupling at 3 years of follow up and the x-axis displays the range of covariate values with the gray area representing a 95% confidence interval for the effect estimate at a given x-axis value. Relationships between continuous covariates and probability of RV-PA uncoupling (the shape of the curves) are similar to the relationships between continuous variables and probability of RV dysfunction (see Figure 3).

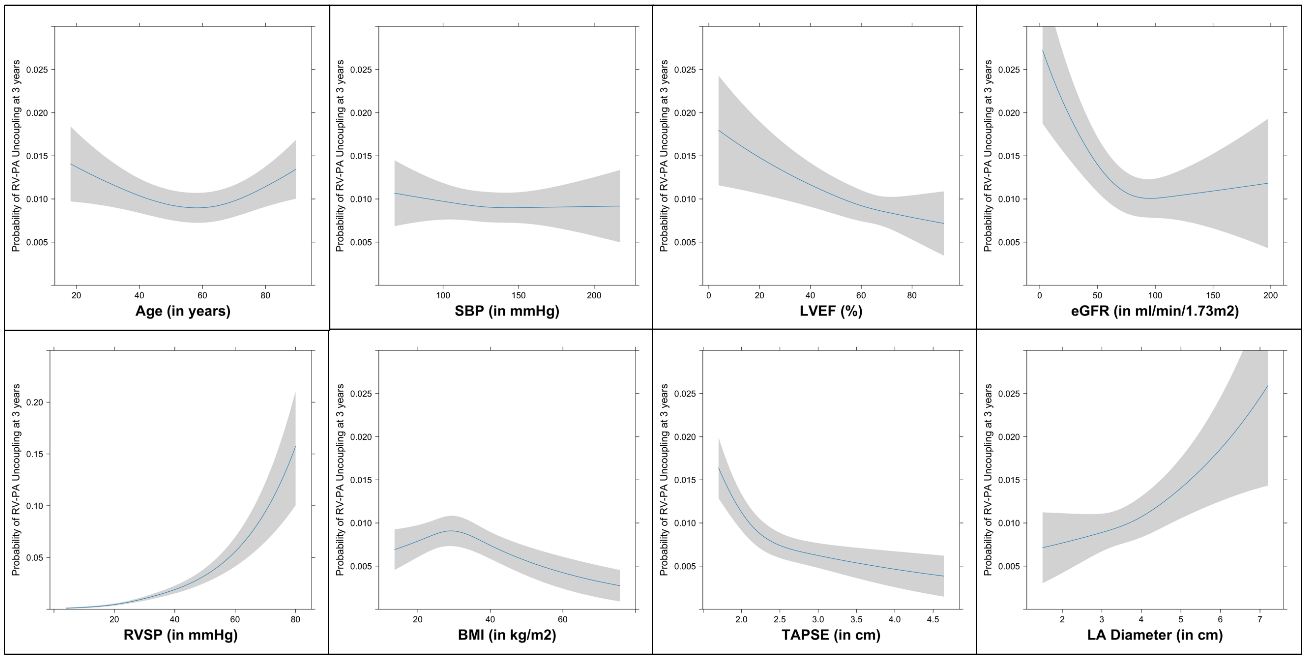
